# Comparing human and AI performance in medical machine learning: An open-source Python library for the statistical analysis of reader study data

**DOI:** 10.1101/2022.05.06.22274773

**Authors:** Scott Mayer McKinney

## Abstract

In seeking to understand the potential effects of artificial intelligence (AI) on the practice of diagnostic medicine, many investigations involve collecting interpretations from several human experts on a common set of cases. In an effort to standardize the process of analyzing the data emerging from such studies, we have released an open-source Python library to perform applicable statistical procedures. The software implements the industry-standard Obuchowski-Rockette-Hillis (ORH) method for multi-reader multi-case (MRMC) studies. The tools can be used to compare a standalone algorithm against a panel of readers, or compare readers operating in two modalities (for example, with and without algorithmic assistance). The software supports both nonequivalence and noninferiority tests. Functions are also provided to simulate reader and model scores, useful for Monte Carlo power analysis. The code is publicly available in our Gitub repository at https://github.com/Google-Health/google-health/tree/master/analysis.

## Background

When we develop machine learning models for interpreting healthcare data, we often want to understand how the models’ predictions compare to those of human experts who usually perform this function. Alternatively, we may wish to evaluate whether predictions of machine learning models can empirically influence human judgment in a favorable way. This is often accomplished by collecting interpretations in a controlled laboratory setting. In these so-called “reader studies”, a panel of experts—such as radiologists, pathologists, or ophthalmologists—independently inspect a set of cases and record their impressions on some quantitative scale (whether continuous, ordinal, or binary). Sometimes, there is an accompanying phase in which the readers view the cases with the assistance of artificial intelligence (AI)-based software. The order of these two phases may be randomized, and they usually occur after a “washout period” of weeks or months, ample time for them to forget the particulars of the case [1].

We then seek to determine how well the algorithm makes diagnostic judgments or follow-up recommendations relative to the experts. For example, can a computer vision algorithm spot breast cancer as well as an average mammographer [2]? On the other hand, we may also wish to assess how the readers are affected by the presence of a decision support tool powered by artificial intelligence. For example, do pathologists grade prostate biopsies more accurately when presented with annotations surrounding abnormal cells [3]?

The literature on how to draw statistical conclusions from multi-reader multi-case (MRMC) studies is well developed, and consensus has gathered around a family of related methods [4–7]. Some open-source tools exist, but they consist largely of R interfaces [8–10], variously backed Java [11] or Fortran [12] code.

However, the majority of modern machine learning and data science today takes place in Python [13], and it’s often convenient to conduct analysis in the same environment in which models are trained and evaluated. Libraries in other languages can be difficult to integrate with Python-based workflows and metrics. Furthermore, since machine learning practitioners are often most fluent in Python, they may have difficulty interpreting, testing, and debugging statistical analysis code in other languages.

To address this gap, we are releasing a Python library to simulate and analyze reader study data. We have used this code in several published papers [2,3,14–16] and are pleased to share it with the broader research community. Complete with unit tests, the code is now available in our Github repository [17]. We hope this will promote a rigorous and standardized approach to evaluating AI-based diagnostic tools.

## Methodology

We provide a suite of functions consisting of two categories, one for *simulation*—the generation of random, synthetic data —and one for *analysis*—the measurement of statistical trends within a given data set. We discuss each in turn.

Table 1 gives an overview of the API.

**Table 1.** Core functions of our Python library for the simulation and analysis of MRMC data under two study designs.

| study design | simulation function | analysis function |
| --- | --- | --- |
| standalone evaluation | `simulate_model_vs_readers` | `model_vs_readers_orh` |
| assisted read | `simulate_dual_modality` | `dual_modality_orh` |

Our implementation is guided in large part by the excellent textbook of Chakraborty [7], to which we owe our gratitude.

### Simulation

To plan a study, it can be useful to simulate data with the same general characteristics as would be expected to emerge from your reader study. For example, simulation can be used to carry out power analysis. (*Power analysis* refers to the process of determining the probability that a study will obtain a statistically significant result, assuming an underlying data distribution, including the noise level and the true effect size. Power can be considered to identify the prudent number of readers and cases to include to best position a study for success.) In a Monte Carlo approach, one can estimate the power of a study by generating many random data sets and empirically measuring how often statistical significance is achieved.

We provide functions to simulate reader scores for two types of experiment designs. Both are based on standard methods [7,18] inspired by the classic model of Roe and Metz [19]. To compare a standalone machine learning model with human performance, we simulate a model score and companion scores from a panel of readers. For an assisted read study, we can simulate pairs of reader scores associated with two different modalities. In either scenario, it is assumed that there is a known binary ground truth status associated with each case. The scores are conditioned on these disease labels and the expected task performance among the different parties. For a discussion of how these quantities are parameterized, see the more recent work by Hillis [20].

Both functions produce normally distributed suspicion scores with a correlation structure that reflects the study design and the nature of the task. In particular, scores assigned to the same case tend to be correlated. Although these human-generated scores are not limited to the traditional range of [0, 100], many performance metrics, like area under the receiver operating characteristic curve (AUC-ROC), are insensitive to absolute scale and merely reflect the ranking of cases [21]. Note that these scores can be post-processed into ordinal or binary judgments (through thresholding, for example) if that is what is expected from the study [22]. For use in power calculations, the many required runs of simulation and analysis may be parallelized with one’s favorite framework.

### Analysis

We provide an implementation of the Obuchowski-Rockette-Hillis (ORH) procedure for analyzing MRMC study data. It was originally formulated for comparing the quality of reads across two modalities: these may be different imaging techniques (e.g. film or digital X-rays) or interpretation modes (e.g. with or without computer assistance). Subsequently, the method was adapted to compare the performance of a standalone algorithm with that of a set of readers [7].

These functions work with an arbitrary metric (or “figure of merit”, in the nomenclature of [7]) defined on the set of suspicion scores and ground truth labels. Common choices are AUC-ROC or classification accuracy, both available in the module `sklearn.metrics`. Sensitivity or specificity can be analyzed by using the accuracy metric and limiting analysis to the positives or negatives, respectively. More exotic choices, like partial [1] or parametric [23,24] AUC-ROC are attainable by providing the right functions. If your metric requires additional inputs beyond ground truth values and scalar suspicion scores, we provide a separate interface to handle this situation.

Regardless of study design, both analysis functions produce an estimate of the study’s effect size, with an associated confidence interval at the specified coverage level (defaulting to the commonly used 95%). To more explicitly address the comparison of interest, the analysis functions produce a *p*-value reflecting the statistical significance of the observed difference between the two modalities, or between the model and average reader. The primary comparison can be aimed at demonstrating nonequivalence (two-sided) or noninferiority (one-sided) [25]. In a nonequivalence study (the default in our software), the null hypothesis states that the average performance between the two groups is equal; under the alternative hypothesis, one is superior to the other. In a noninferiority study, the null hypothesis holds that one particular group is inferior to the other by at least some pre-specified positive margin *δ > 0*. (The margin should be a clinically meaningful quantity chosen *a priori*, ideally before the study is conducted and certainly before the test data is inspected.) If the null hypothesis is rejected, we can claim that the performance of the second modality (or model) is not appreciably worse than the first modality (or average reader). See Figure 1. To assess statistical significance of the study, the *p*-value is commonly compared against an “alpha level” of 0.05, but this threshold may need to be adjusted to account for multiple comparisons [26].

**Figure 1.**
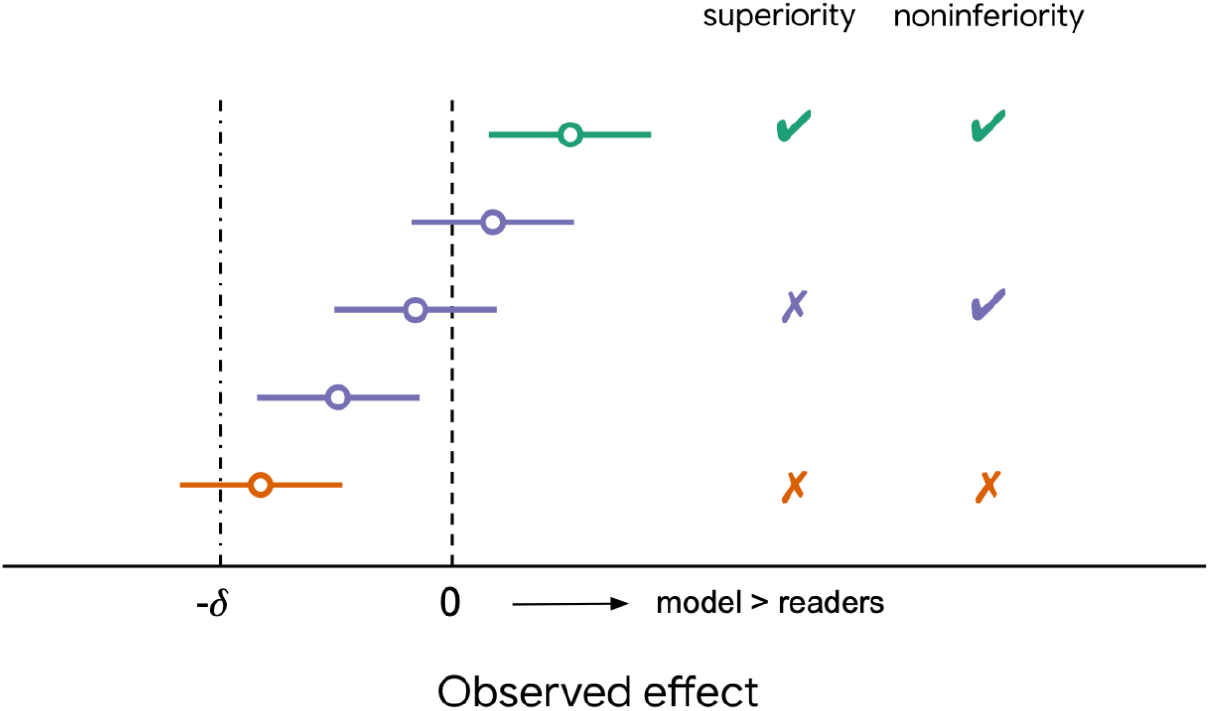
Various outcomes of a reader study. An effect size of 0 corresponds to no difference between model and readers (or the two arms of a dual modality study). The parameter *δ* is a positive, clinically significant margin chosen *a priori*. Each horizontal colored line denotes the 95% confidence interval (CI) surrounding a point estimate, indicated by a circle. If the lower bound of the CI exceeds 0, superiority is demonstrated. If the lower bound exceeds -*δ*, noninferiority is demonstrated. See [25] for an extended discussion.

Where equivalent functions were available, our Python code was benchmarked against the results produced by the RJafroc package [9].

## Limitations

Many extensions could add versatility and flexibility to this library. The present version of the software only supports vanilla reader study designs: it assumes a fully crossed or factorial setup in which each case is read by every reader. It does not gracefully handle missing data (data imputation is left to the user), nor does it handle more exotic situations like the “split-plot” design, though it may be straightforward to do so [27]. Finally, only two arms are supported, though a reader study might have several. For example, one might wish to compare a model against two subsets of readers with different levels of training, or to compare several manifestations of a decision support tool with different user experience choices. In these situations, users may employ the provided functions to pursue a strategy of pairwise comparison.

The simulation paradigm is sensitive to the variance parameters, which should be carefully calibrated to the discrimination task of interest. This cannot be done reliably in the absence of real pilot data. Even with such data, the methodology for calibrating the parameters is not straightforward [7]. For the dual modality scenario, we leverage presets which may not apply well to all tasks [20].

## Sample usage

### Standalone evaluation

The following block of code simulates and analyzes the scores from one model and 8 readers on 300 cases, 100 of which are positive for some disease of interest. On the binary classification task of separating healthy cases from those with evidence of disease, the model is expected to have an AUC-ROC of 0.9, while the readers have, on average, an AUC-ROC of (The variance parameters `sigma_r` and `sigma_c` were chosen to yield a performance spread typical of radiology studies, but we did not invoke any particularly principled procedure.) The results of the simulation are arrays of size (300,) and (300, 8) for the model and readers, respectively. The analysis step returns a `TestResult` object containing the statistical quantities of interest. The corresponding ROC curves are shown in Figure 2a. In this instantiation of the data, a difference of 0.106 AUC was observed, with associated 95% confidence interval of (0.064, 0.147). The two-sided *p*-value for nonequivalence was 2.4 × 10^−4^, demonstrating a clear statistical difference. Since the observed difference is positive, we conclude that the model is superior.

**Figure 2.**
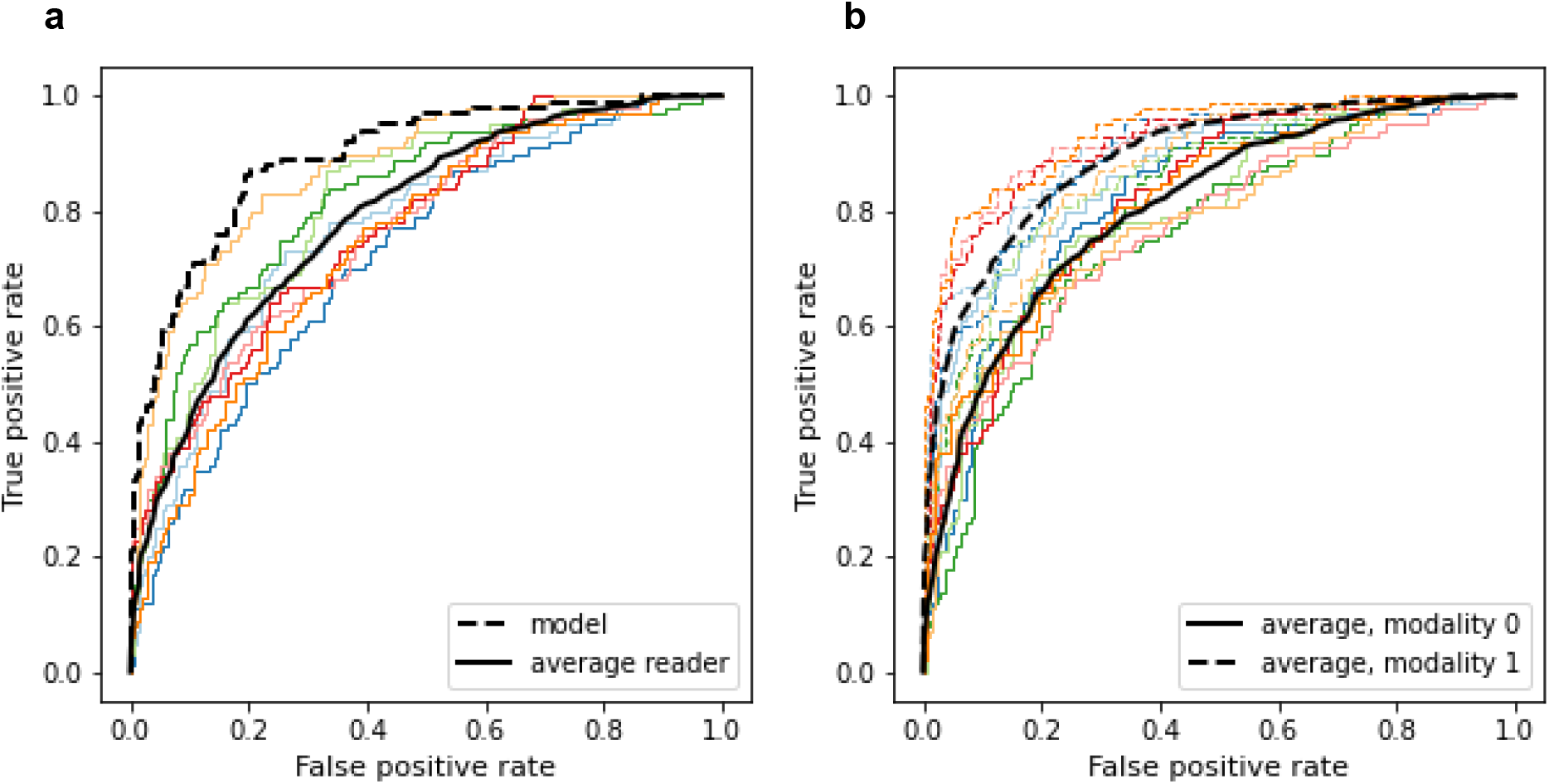
Simulated receiver-operating characteristic (ROC) curves of 8 readers on *n* = 300 cases, 100 of which are positive. (**a**) Data that might be observed in a standalone evaluation study. The model (dashed curve) has an expected area under the curve (AUC) of 0.9, while the average reader has an expected AUC of 0.8. (**b**) Data that might be observed in a dual-modality assisted read study. There are two curves for each reader with the same color but different line style. Solid curves represent readers operating in the first modality (with average AUC of 0.8), while dashed curves represent readers operating in the second modality (with average AUC of 0.9).

~~~
import numpy as np
import sklearn.metrics
import reader_study

num_positives = 100
num_negatives = 200
disease = num_positives * [1] + num_negatives * [0]
num_readers = 8

model_score, reader_scores = reader_study.simulate_model_vs_readers(disease,
    model_auc=0.9,
    reader_auc=0.8,
    sigma_r=0.1,
    sigma_c=0.6,
    num_readers=num_readers,
    rng=np.random.RandomState(2022))

reader_study.model_vs_readers_orh(disease,
                                 model_score,
                                 reader_scores,
                                 fom_fn=sklearn.metrics.roc_auc_score)
~~~

### Dual modality study

The following block of code simulates and analyzes the suspicion scores of 8 readers participating in an MRMC study with 300 cases (100 of which are again positive) and two arms or modalities. These modalities might correspond to interpretation with and without the assistance of a computer-aided detection (CAD) device. In this simulation, the second arm has a higher median AUC-ROC of 0.9 compared to 0.8 in the first. The returned array of reader scores has size (300, 8, 2). The pairs of reader ROC curves, along with the averages from each arm, are shown in Figure 2b. This run produced an effect size of 0.086 with a 95% confidence interval of (0.053, 0.12). The two-sided *p*-value for nonequivalence was 2.0 × 10^−5^, demonstrating a clear statistical difference. Since the observed difference is positive, we conclude that the second modality is superior to the first.

~~~
import numpy as np
import sklearn.metrics
import reader_study

num_positives = 100
num_negatives = 200
disease = num_positives * [1] + num_negatives * [0]
num_readers = 8

_, scores = reader_study.simulate_dual_modality(disease,
                                                modality_0_auc=0.8,
                                                modality_1_auc=0.9,
                                                structure=‘LL’,
                                                num_readers=num_readers,
                                                rng=np.random.RandomState(2021))

reader_study.dual_modality_orh(disease,
                              scores,
                              fom_fn=sklearn.metrics.roc_auc_score,
                              verbose=False)
~~~

## Data Availability

All of the associated code is publicly available on Github.

https://github.com/Google-Health/google-health/tree/master/analysis

## Glossary

- *reader*: a human annotator, usually an expert trained to interpret medical signals or images and identify signs of pathology.
- *case*: a single unit of analysis in a reader study. It usually corresponds to the data (e.g. a collection of images) from one patient, associated with a known disease status, which readers attempt to assess.
- *modality*: one of typically two arms in a multi-reader multi-case study. Example modality pairs are (CT, MRI); (film X-ray, digital X-ray); (fundus imaging, OCT); (assisted read, unassisted read).
- *assisted read study*: a multi-reader multi-case study in which readers interpret cases with and without computer assistance (e.g. from a machine learning algorithm). The two reading conditions constitute different “modalities.”
- *standalone evaluation*: a study in which the output of a machine learning algorithm is compared to the judgments from a panel of readers.
- *figure of merit:* any valid measure of performance, usually defined on the set of suspicion scores and ground truth labels. Examples are AUC-ROC and accuracy.

## Acknowledgments

We appreciate Rory Sayres and Ellery Wulczyn for reviewing and testing the code. Thanks also to Yun Liu and Christopher Kelly for detailed feedback on this manuscript.

